# Post-operative pain management practice and associated factors among nurses working at public hospitals, in Oromia region, Ethiopia

**DOI:** 10.1101/2022.04.14.22273889

**Authors:** Abebe Dechasa, Abdo Kurke, Desalegn Abdisa, Yonas Gurmu

## Abstract

**Background:** Management of postoperative pain leads to positive patient progress and shortens the duration of hospital stay. Nurses, who are the majority in almost all hospitals and spend most of their time with the patients, are expected to play big role in the postoperative pain management practice. However, there is paucity of information regarding postoperative pain management practice and its associated factors among nurses.

**Objective:** To assess postoperative pain management practice and associated factors among nurses working at public hospitals, in Oromia Region, Ethiopia, 2020.

**Methods:** Institutional based cross sectional study was employed among randomly selected 377 nurses working at public hospitals in Oromia region, Ethiopia. Data was collected by distributing structured self-administered questionnaires that adapted from different literatures. The data were entered into Epi data version 3.1 and exported to SPSS version 22 for analysis. Variables with significant association in the bivariate analyses were entered into a multivariable regression analysis to identify the independent factors associated with nurses’ postoperative pain management practice. Significant factors were declared at P<0.05.

**Result:** The result showed that, 66% of nurses had good pain management practice. Nurses favorable attitude towards post-operative pain management [AOR: 4.698, 95% CI: (2.725-8.100)], having access to read pain management guideline [AOR: 3.112, 95% CI: (1.652-5.862)], adequate knowledge of post-operative pain management [AOR: 2.939, 95% CI: (1.652-5.227)], working at Operation Room [AOR: 2.934, 95% CI: (1.27-6.795)] and received training on pain management [AOR: 3.289, 95% CI: (1.461-7.403)] were significantly associated with the practices of postoperative pain management.

**Conclusion and recommendation:** Sixty six percent of participants (nurses) have a good level of practice of postoperative pain management. Training on post-operative pain management (POPM), access to pain management guidelines, knowledge and attitude are significant factors in post-operative pain management practice. Governmental and other bodies concerned to post-operative care quality needs to show commitment on availing infrastructures like pain management guideline and improving nurses knowledge and attitude.

## Introduction

Post-operative pain (POP) is a form of acute pain following surgical trauma (1) as a result of the inflammatory reaction and initiation of afferent neurological barriers. The pain is felt in response to the inflammatory process resulted from tissue injury during surgical procedure like skin incision, tissue dissection, manipulation and traction (2). Unless adequately managed post-operative pain management can be complicated to delayed ambulation, reduced patient satisfaction and increased incidence of pulmonary complication(3).

Currently, it was estimated that about 28-32% of global disease requires surgical intervention (4). Over five million surgical interventions are needed in Ethiopia each year (5). The rise in the number of operations is not without risk. For instance, in the United States, between 10% and 60% and in Ethiopia 22% of patients were developed chronic pains as results of poorly managed postoperative pain (6, 7). Furthermore poorly managed postoperative pain can have a negative impact and is always associated with delayed mobility which can lead to delayed wound healing, deep vein thrombosis, pulmonary complications secondary to suppression of effective coughing, anxiety, sleep disturbance, myocardial infarction, depressed immune function and also can progress to chronic pain which impairs the ability to carry out daily activities and finally may leads to diminished quality of life (8).

Since, pain relief has been recognized as a human right and also considered as the “fifth” vital sign that must be regularly assessed and managed, the nurses should give inordinate attention to control postoperative pain(9). The roles and responsibilities of nurses in pain management; according to American nurses association (ANA), includes, assessment of pain, plan for pain management strategies and evaluation of responses of the patients for the given interventions and to take actions accordingly. Since, nurses are always spending 24 hours at bedside to give care for the patients and also they are the point of contact between other health professionals and the patients, they are expected to play vital role in postoperative pain management practice (10).

Post-operative pain management practice is a set of activities and an important aspect of nursing care to alleviate pain for the patients by pharmacologic and non-pharmacologic methods. These activities include assessing the pain, providing appropriate interventions to relieve the pain and reassessing the patients’ pain after intervention. Assessing pain is the first and crucial step for properly managing pain. Techniques for pain assessments include patients self-report and observing for patients’ physiological and behavioral responses to pain. The self-reporting methods include numeric rating scale (NRS), verbal rating scale (VRS), visual analogue scale (VAS) and the faces pain scale (FPS) (11).

Although, postoperative pain management continues to be a problem in developed and developing countries, sadly the suffering from untreated postoperative pain is larger and more worrying among the economically disadvantageous individuals in developing countries. Alleviating patients suffering is a core ethical and legal obligation for health professionals. However, in Ethiopia, discomfort due to post-operative pain remains prevalent and affects between 47%-100% of patients after surgery (7).

Undertreated POP can contribute to different socio economic burden such as increasing health care cost by delaying discharge directly and indirectly as a result of absenteeism from job or loss of production. Nowadays, there is a growing awareness on the etiology of pain and an advancement of pharmacological and non-pharmacological pain management. Despite of this, patients still experience unacceptable pain after surgical procedures. The study was conducted by aiming to identify nurses post-operative pain management practice at public hospitals found in Oromia, Ethiopia; so that the finding will best serve to prioritize the problem and develop strategies for improving post-operative pain management.

## Methods

### Study area, period and design

The study was done at public hospitals found in West Shoa zone, Oromia regional state, Ethiopia from June 1 to August 30/2020. There are one referral, three general and four districts (total of eight public hospitals) in that zone. These hospitals provide different health service ranging from prevention of disease to surgical therapies for peoples in the area and closer zones in the Oromia region. So that post-operative nursing care is given for patients in need of the services. Cross sectional study design was implemented to identify post-operative pain management practice and associated factors among those nurses.

### Population

#### Source Population

All nurses, who were working in surgical ward, Minor operation room (OR) and Major operation room (OR), Recovery rooms, Emergency, obstetrics and gynecology wards were included.

#### Study Population

All randomly selected nurses working in surgical ward, minor OR, major OR, recovery room, emergency room, obstetrics and gynecology wards were considered the study population.

#### Sample size determination

The sample size was determined by using single population proportion formula by considering that 65.2% nurses had good post-operative pain management practice (12) at a 5% level of significance, 5% margin of error and considering 10% non-response rate. With this calculation the final sample size was 384.

#### Sampling techniques

All public hospitals in the study area were included in the study. The total calculated sample size (384 nurses) was proportionally allocated to each hospital based on the number of their professional nurse. Finally by using the registration number of nurses at each hospital, participants of the study were selected randomly by the lottery methods.

#### Study variables

##### Dependent variable

level of Post-operative pain management practice

##### Independent variables

Socio demographic characteristics (Sex, Age, Marital Status, Educational status, Experience, Working unit/ward), Knowledge towards postoperative pain management, Attitude towards postoperative pain management, organizational factors (Availability of standardized tools, Guideline, Training on pain management).

### Operational definitions

#### Good Practice

Refers to those study participants, who have scored mean and/or above the value of the total 18 practice questions.

#### Poor Practice

Refers to those study participants who have scored below the mean value of the total 18 practice questions.

#### Knowledge

Is measured by fifteen items in yes/no format. Correct answer was given “1” and “0” was given for incorrect and for not sure. Those who scored mean and above were labeled as having adequate knowledge where as those who scored less than mean labeled as having inadequate knowledge about post-operative pain management.

#### Attitude

Is measured by nine items in agree/disagree format. For correctly responded item “1” was given and “0” was given for incorrect and don’t know. Those who scored mean and above considered as having favorable attitude where as those who scored below mean have unfavorable attitude towards postoperative pain management (12-14).

### Data collection tool and data quality control

Structured self-administered questionnaire was used to collect data. The questioners were adapted from different studies conducted previously and modified. To assure the data quality, data collection tool was prepared after the intensive reviewing of relevant literatures and similar studies. The tool was reviewed by expert’s panels. One clinical nurse specialist, three lecturers (masters of Science in nursing) and one registered nurse (BSC nurse) were participated in the panel (review of the questionnaire). The questionnaire was pretested on 5% of the study population at similar health care facilities. Training was given for data collectors by the principal investigator.

### Data processing and analysis

The collected data was coded, cleaned, and entered in to Epi Data version 3.1software and finally exported to statistical package for social study (SPSS) version 22 Software for analysis. Descriptive analyses were performed first to understand the general characteristics of all the study variables. The results were presented in tables and graphs using summary measures such as percentages and mean. Bivariate logistic regression was carried out to identify the factors associated with nurses’ postoperative pain management practice. Hosmer-Lemeshow test was done to test for model fitness, the result was 0.45. Variables with p < 0.25 in the bivariate analyses were entered into a multivariable logistic regression analysis to identify the independent factors associated with the outcome variable. Finally, the result of bivariate and multivariable logistic regression analysis was presented in a crude odds ratio (COR) and adjusted odds ratio (AOR) with 95% confidence intervals. P ≤ 0.05 was considered statistically significant.

## Result

### Socio-demographic characteristics

A total of 384 questionnaires were distributed, of which 377 were completed and returned with the response rate of 98.2%. The majority of participants, 227(60.2%) were male, 200(53.1%) were married and 240 (63.7%) were between the age group of 26 and 34 years (**Table 1**).

**Table 1:** Socio-demographic characteristics of respondents, working at public hospitals in Oromia region, Ethiopia, 2020

| Variables | Category | Frequency(n=377) | Percentage (%) |
| --- | --- | --- | --- |
| Sex | Male | 227 | 60.2 |
|  | Female | 150 | 39.8 |
| Age | < 25 | 71 | 18.8 |
|  | 26-34 | 240 | 63.7 |
|  | >35 | 66 | 17.5 |
| Ethnicity | Oromo | 360 | 95.5 |
|  | Amhara | 17 | 4.5 |
| Marital status | Married | 200 | 53.1 |
|  | Single | 177 | 46.9 |
| Religion | Protestant | 217 | 57.6 |
|  | Orthodox | 103 | 27.3 |
|  | Muslims | 40 | 10.6 |
|  | Wakefata | 17 | 4.5 |
| Educational level | Diploma | 28 | 7.4 |
|  | Bachelor degree | 346 | 91.8 |
|  | Masters | 3 | 0.8 |
| Years of experience | <5 | 234 | 62.1 |
|  | 6-9 | 75 | 19.9 |
|  | >10 | 68 | 18 |
| Work experience in surgical unit (in years) | <1 | 199 | 52.8 |
|  | 2-4 | 140 | 37.1 |
|  | >5 | 38 | 10.1 |
| Current area of practice | Medical ward | 65 | 17.2 |
|  | Emergence ward | 70 | 18.6 |
|  | Ob/Gyne ward | 65 | 17.2 |
|  | OR and Recovery | 72 | 19.1 |
|  | Surgical ward | 105 | 27.9 |

### Knowledge of nurses towards post-operative pain management

The mean score for knowledge was 8.89 with standard deviation of (±2.85). Thus, the results revealed that, from the total of 377 study participants, about 54.9 % (95% CI: (50.1, 60.2)) had adequate knowledge about POP management (**figure 1**).

**Figure 1:**
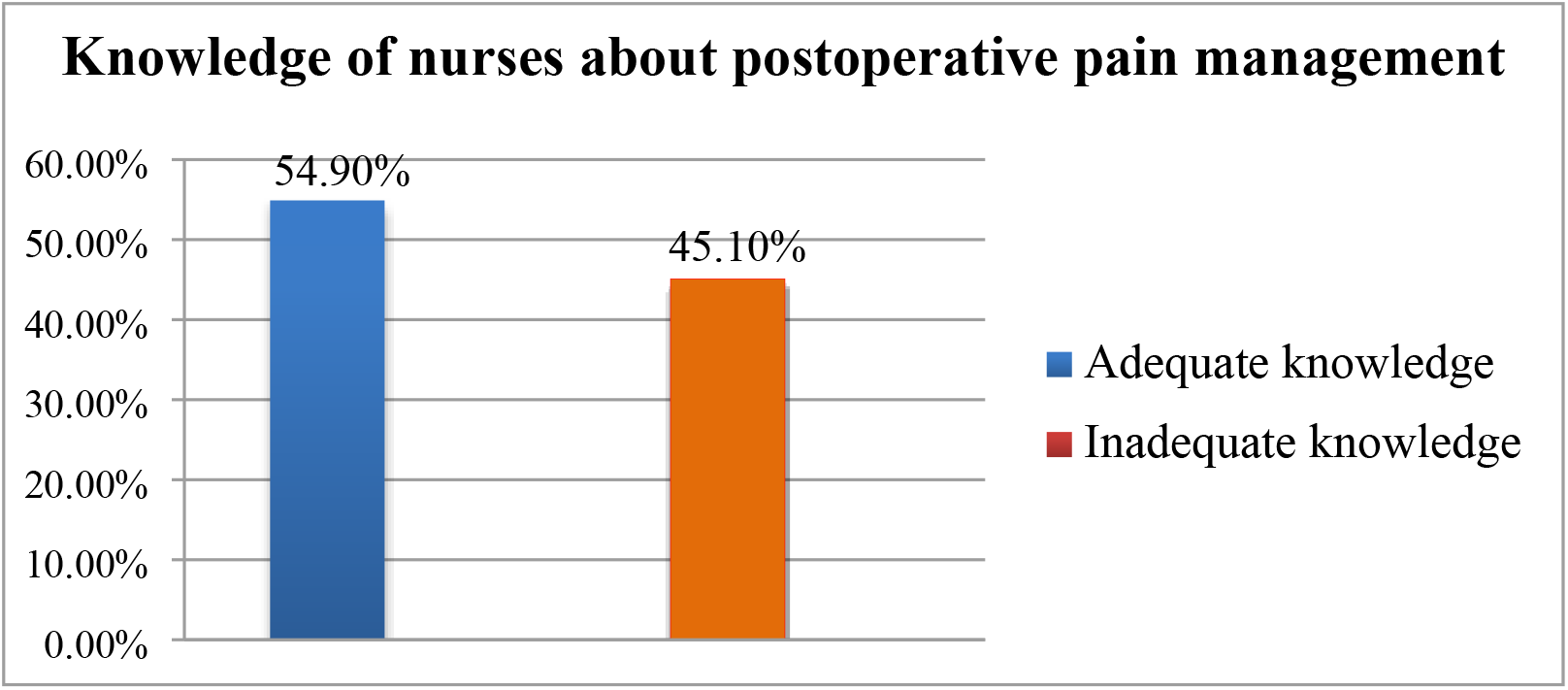
Knowledge levels of nurses on postoperative pain management at public hospitals in Oromia region, Ethiopia, 2020.

### Nurses’ attitude of postoperative pain management

The mean score for attitude was computed and it was 4.99 with standard deviation of 1.73. According to the classification outlined in the operational definition the percentage score of the categories showed that, among 377 respondents, 59.4% (95% CI: (54.6, 64.5)) of participants had favorable attitude towards post-operative pain management practice (**figure 2**).

**Figure 2:**
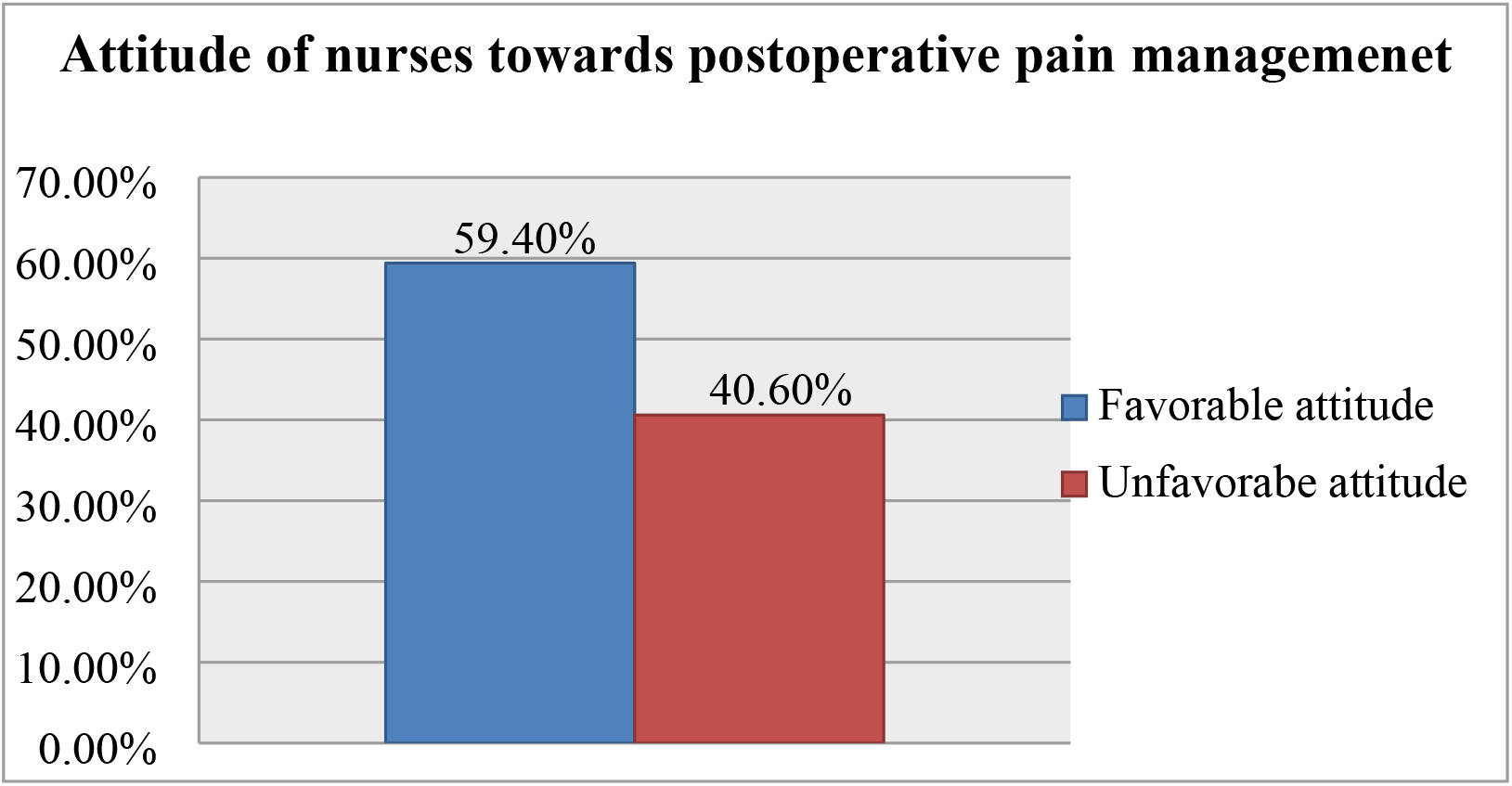
Attitude levels of nurses towards postoperative pain management at public hospitals in west shoa zone, Ethiopia, 2020.

### Practices of nurses on postoperative pain management

The responses of nurses to the nine practice questions are computed and dichotomized in to good practice and poor practice. The mean score of the self-report practice of post-operative pain management was 10.37 with standard deviation of (±2.89). It was calculated based on the category specified in the operational definitions. Accordingly this study revealed that, about two third (66%) (95% CI: (61, 71) of the respondents had good postoperative pain management practice **(Table 2)**.

**Table 2:** Practices of nurses on postoperative pain management, working at public hospitals in Oromia region, Ethiopia, 2020

| Variables | Yes N (%) | No N (%) |
| --- | --- | --- |
| Do you assess pain for the patients those able to communicate? | 327(86.7%) | 50 (13.3%) |
| Do you encourage the use of transcutaneous electrical nerve stimulator for pain management? | 4(1%) | 373(99%) |
| Do you combine opioids with NSAID's rather than single analgesic agents when managing POP as suggested by World health organization? | 302(80.1%) | 75 (19.9%) |
| Do you document the findings after pain assessment? | 110(29.2%) | 267 (70.8%) |
| Do you encourage prayer by patients or religious leader postoperatively? | 206(54.6) | 171 (45.4%) |
| Do you administer ordered pain medication, around the clock (regularly) as ordered? | 374(99.2%) | 3(0.8%) |
| Do you use music therapy to reduce pain? | 2(0.5%) | 375 (99.5%) |
| Do you reassess pain after giving pain medication in order to evaluate the effectiveness of pain medication? | 338(89.7%) | 39 (10.3%) |
| After surgery, do you provide comfortable positions to help relieve pain? | 360(95.5%) | 17 (4.5%) |
| Do you ask and help to support the painful areas when moving or coughing after surgery? | 283(75.1%) | 94 (24.9%) |
| Do you provide clean, calm and ventilated ward environment for postoperative pain management? | 259(68.7%) | 118 (31.3%) |
| Do you lay patients on neat, well-laid bed postoperatively? | 294(78%) | 83 (22%) |
| Do you use massage and stretch to reduce postoperative | 283(75.1%) | 94 (24.9%) |
| pain? |  |  |
| Do you apply heat and cold compresses to manage POP? | 288(76.4%) | 89 (23.7%) |
| Do you encourage early ambulation/exercise with analgesia? | 347(92%) | 30 (8%) |
| Do you encourage use of acupuncture? | 2(0.5%) | 375(99.5%) |
| Do you use patient distraction, relaxation, and guided imagery postoperatively to reduce pain? | 131(34.2%) | 246(65.2%) |
| Do you usually dress, bandage, splint and reinforce wound sites postoperatively? | 359(95.2%) | 18(4.8%) |

### Organizational related factors

According to the nurses response regarding the organizational factors majority, 273(72.4%) of the participants reported that they have not taken any training regarding postoperative pain management while 221(58.3%) did not accessed post-operative pain management guidelines to use for practice. Among those received training regarding postoperative pain management 59(56.7%), 39(37.5%), 2(1.9%) and 4(3.8%) received training by the means of lecturing, course, conference and work shop respectively.

### Factors associated with postoperative pain management practice

To assess the factors associated with the nurses’ postoperative pain management practice, bivariate analysis was done first. Accordingly, ten of the variables age of the participants, marital status, level of education, work experience, experience in postoperative area, current area of practice, training related to pain management, access to read pain management guideline, knowledge and attitude of the participants regarding POP management were found to be significantly associated with the nurses ‘POP management practice at p-value of 0.25. These variables were included in multiple logistic regressions analysis. The model fit was checked by Hosmer and Lemeshow test (p-value=0.45) and it was fitted.

After adjustment, attitude, getting access to read guidelines, training, knowledge and current area of practice were significantly associated with the nurses’ postoperative pain management practice. Accordingly, respondents who had Favorable attitude were almost 5 times more likely to practice than those who had unfavorable attitude [AOR: 4.698, 95% CI: (2.725, 8.100)]. Respondents who have taken POP management training were 3.2 times more likely to practice than those who did not take such training [AOR: 3.289, 95% CI: (1.461, 7.403)]. Similarly, study participants who get access to read pain management guidelines were 3.1 times more likely to practice compared to their counterparts [AOR: 3.112, 95% CI: (1.652, 5.862)]. The study also revealed that respondents who had adequate knowledge on postoperative pain management were 2.9 times more likely to practice than those who had inadequate knowledge [AOR: 2.939, 95% CI: (1.652, 5.227)] and participants those who were currently practicing in Operation Room were 2.9 times more likely practice compared with those practicing in medical ward [AOR: 2.934, 95% CI: 1.267, 6.795 P<0.012] (**Table 3**).

**Table 3:**
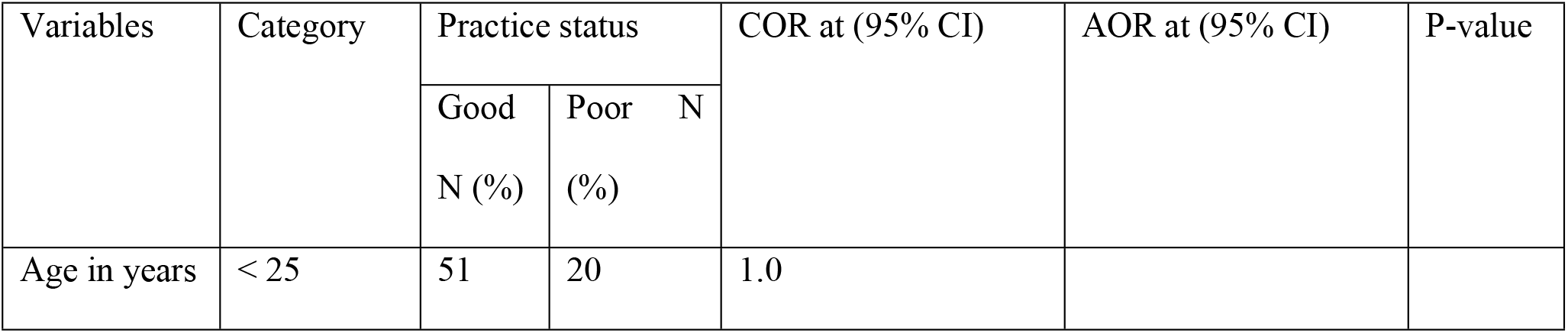
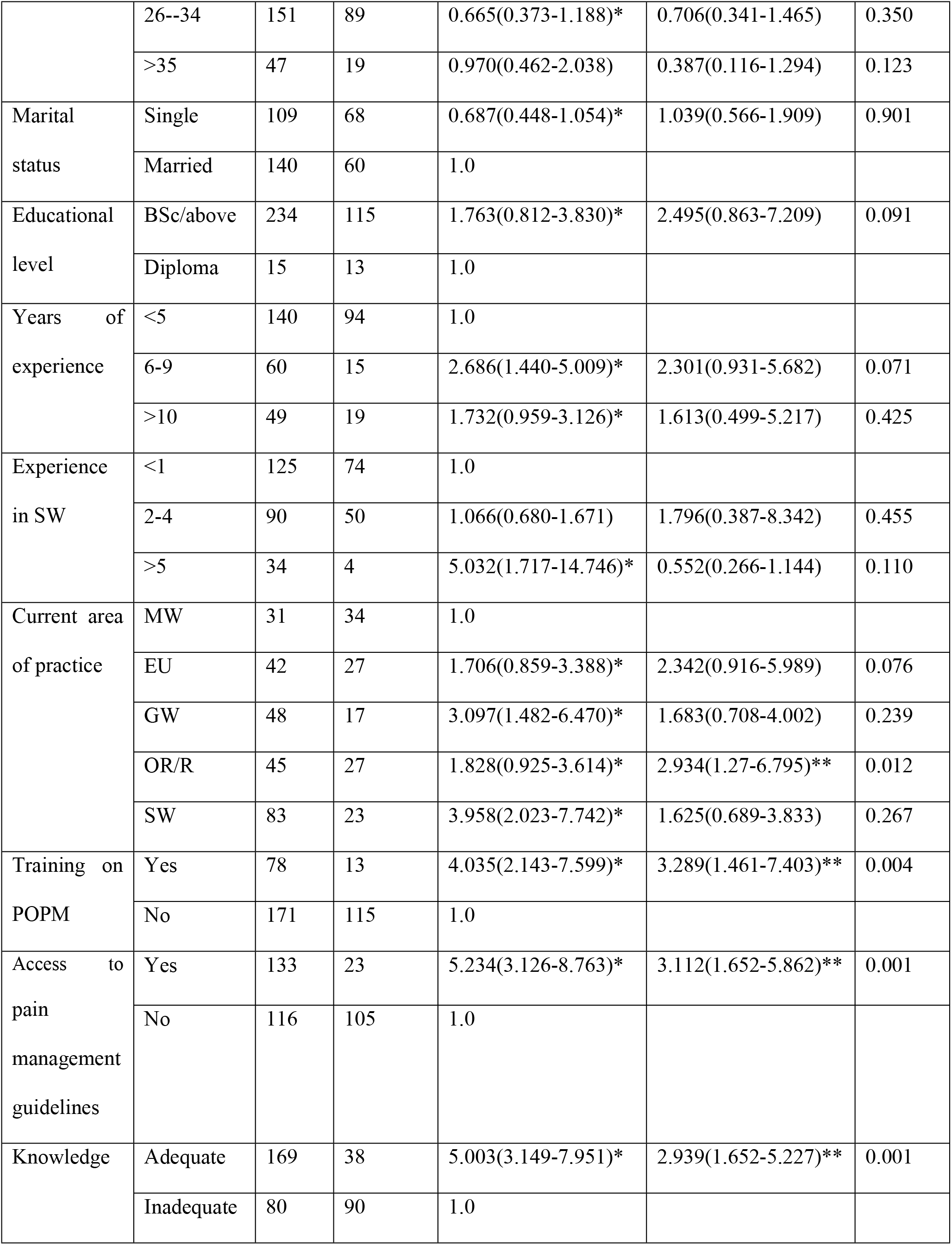
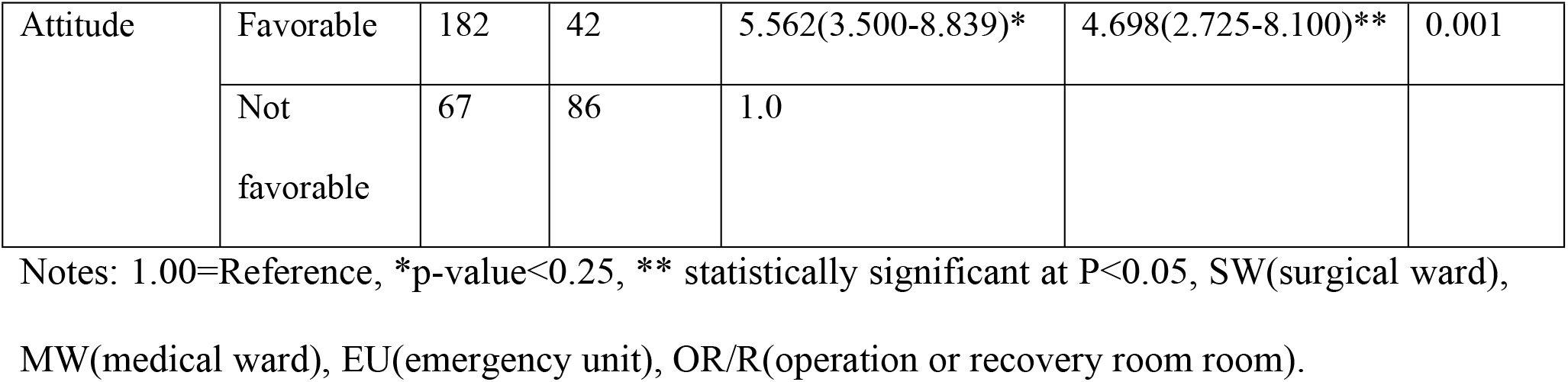
Binary and multiple logistic regression analysis results on factors associated with postoperative pain management practice among nurses working at public hospitals in west shoa zone, Ethiopia, 2020.

## Discussion

The current study revealed that overall postoperative pain management practices among 66%, (95% CI: 61, 71) nurses was found to be good. This finding is lower as compared to the study conducted in Rwanda on postoperative pain management, which was 88% (15). However, the finding of this study was greater than the study conducted in Addis Ababa in which only 6% of them had had good practice and the study done in Arsi zone, southeastern Ethiopia, where nearly half (47.9%) of their study participants had good pain management practice(12,16). This discrepancy may be due to nursing workload, access to read guideline, sample size, and use of different data collection tools. This study revealed that those who had favorable attitude were nearly five times more likely to have good postoperative pain management practices than those who did not. This is consistent with a study conducted in Addis Ababa and Ghana (13, 16).

Those who had received training were more than three times more likely to have good pain management practices than those who didn’t. This finding was comparable with the study conducted at Debra Berhan, Northern Ethiopia (17). This is because those individuals who had taken POP management training could have current information on pain management which can promote the practice. The current study also identified that, those who got access to read pain management guidelines were 3.1 times more likely practices than their counterparts. This finding is supported by studies conducted in Greece and Debra Berhan (17, 18). This is because accessibility to refer guidelines can enhance the practices of POP management, according to the recommended standard. It is also currently the most advisable for clinicians that keeping up-to-date with evidence-based practice.

Individuals who were knowledgeable were nearly three (2.9) times more likely to have good practice than those who had inadequate knowledge. This finding was in line with the finding from Rwanda and Arsi zone of southeastern Ethiopia (12, 15). The possible justification is that, the right knowledge about pain and its management practice can avoids confusion regarding POP and the disease condition, which can also create clear understanding of its negative impact on the patients and on health institutions, unless appropriately managed. This study also showed association between nurses’ current working area and level of practice.

### Conclusion and recommendation

Sixty six percent of participants (nurses) have a good level of practice of postoperative pain management. Training on post-operative pain management (POPM), access to pain management guidelines, knowledge and attitude are significant factors in post-operative pain management practice. Regional health bureau, Zonal health offices, hospital administrations and other concerned bodies needs to work for enhancing post-operative pain management through organizing different trainings to improve knowledge and attitude of nurses and timely distributing standard pain assessment and management guidelines for enhancing accessibility.

## Data Availability

All relevant data are within the manuscript

## Abbreviations

OR: operation room
POP: post-operative pain
SD: Standard deviation
SPSS: Statistical Package for the Social Sciences

## Availability of data

Datasets used are available from the corresponding authors on reasonable request.

## Ethical and legal Consideration

Ethical clearance was obtained from research Ethics Review committee of Ambo University, college of medicine and health science. Official letter of cooperation was written from Ambo University to all public hospitals in the study area. Participants of the study were being informed that; their participation is voluntary based, the procedures of the data collection, confidentiality of the collected data, and potential risk and benefits related with participation.

## Acknowledgments

We would like to thank all zonal health department and hospital administrative in west shoa zone of Oromia regional state and all study participants for their cooperation.

## Author Contributions

Abebe Dechasa, Abdo Kurke and Desalegn Abdisa were involved in the selection of design, development of the research proposal, data analysis, writing up of the different drafts and finalizing the research. Yonas Gurmu has participated in the reviewing of the different drafts of the study and drafting the amnuscript.

## References

1. Ceyhan D, Ms G. Postoperatif ağrı sadece nosiseptif ağrı mıdır. Ağrı. 2010;22(2):47–52.

2. Jungquist CR, Vallerand AH, Sicoutris C, Kwon KN, Polomano RC. Assessing and managing acute pain: a call to action. AJN The American Journal of Nursing. 2017 Mar 1;117(3):S4–11.

3. Meissner W, Huygen F, Neugebauer EA, Osterbrink J, Benhamou D, Betteridge N, Coluzzi F, De Andres J, Fawcett W, Fletcher D, Kalso E. Management of acute pain in the postoperative setting: the importance of quality indicators. Current medical research and opinion. 2018 Jan 2;34(1):187–96.

4. Global Surgery. Global Surgery & Anaesthesia Statistics: The Importance of Data Collection. Harvard, Medical Shool. 2018.

5. WHO. Surgical Care Systems Strengthening. 2017. 1–45 p.

6. Gan TJ. Poorly controlled postoperative pain: prevalence, consequences, and prevention.Journal of pain research. 2017;10:2287.

7. Eshete MT, Baeumler PI, Siebeck M, Tesfaye M, Haileamlak A, Michael GG, Ayele Y, Irnich D. Quality of postoperative pain management in Ethiopia: A prospective longitudinal study. Plos one. 2019 May 1; 14(5):e0215563.

8. Corke P. Postoperative pain management. Australian Prescriber. 2013 Dec;36(6):202–5.

9. Chatchumni M, Namvongprom A, Eriksson H, Mazaheri M. Thai Nurses’ experiences of post-operative pain assessment and its’ influence on pain management decisions. BMC nursing. 2016 Dec;15(1):1–8.

10. Coyne P, Mulvenon C, Paice JA. American Society for Pain Management Nursing and Hospice and Palliative Nurses Association position statement: Pain management at the end of life. Pain Management Nursing. 2018 Feb 1;19(1):3–7.

11. Chou R, Gordon DB, de Leon-Casasola OA, Rosenberg JM, Bickler S, Brennan T, Carter T, Cassidy CL, Chittenden EH, Degenhardt E, Griffith S. Management of Postoperative Pain: a clinical practice guideline from the American pain society, the American Society of Regional Anesthesia and Pain Medicine, and the American Society of Anesthesiologists’ committee on regional anesthesia, executive committee, and administrative council. The journal of pain. 2016 Feb 1;17(2):131–57.

12. Wurjine T, Nigussie B. Knowledge, attitudes and practices of nurses regarding to post-operative pain management at hospitals of Arsi zone, Southeast Ethiopia, 2018. Women’s Health. 2018;7(5):130–5.

13. Menlah A, Garti I, Amoo SA, Atakro CA, Amponsah C, Agyare DF. Knowledge, Attitudes, and Practices of Postoperative Pain Management by Nurses in Selected District Hospitals in Ghana. SAGE Open Nurs. 2018 Nov 9;4:2377960818790383. doi: 10.1177/2377960818790383. PMID: 33415201; PMCID: PMC7774443.

14. Liyew B, Dejen Tilahun A, Habtie Bayu N. Knowledge and Attitude towards Pain Management among Nurses Working at University of Gondar Comprehensive Specialized Hospital, Northwest Ethiopia. Pain Research and Management. 2020; 2020.

15. Umuhoza O, Chironda G, Katende G, Mukeshimana M. Perceived knowledge and practices of nurses regarding immediate post-operative pain management in surgical wards in Rwanda. A descriptive cross-sectional study. International Journal of Africa Nursing Sciences. 2019; 10: 145–51.

16. Mulugeta E. Assessment of Adult Postoperative Pain Management Practice Among Nurses Working in Addis Ababa Public Hospitals, Addis Ababa, Ethiopia, 2015.

17. Dessie M, Asichale A, Belayneh T, Enyew H, Hailekiros A. Knowledge and Attitudes of Ethiopian Nursing Staff Regarding Post-Operative Pain Management: A Cross-Sectional Multicenter Study. Patient Relate Outcome Meas. 2019;10:395-403. Published 2019 Dec 23. doi:10.2147/PROM.S234521.

18. Kiekkas P, Gardeli P, Bakalis N, Stefanopoulos N, Adamopoulou K, Avdulla C, Tzourala G, Konstantinou E. Predictors of nurses’ knowledge and attitudes toward postoperative pain in Greece. Pain Manag Nurs. 2015 Feb;16(1):2–10. doi: 10.1016/j.pmn.2014.02.002. Epub 2014 Jun 26. PMID: 24981120.

